# SHANK3-anchored reverse phenotyping identifies a rare-variant-enriched cognitive-motor subgroup of autism

**DOI:** 10.64898/2026.05.07.26352644

**Authors:** Aishwaryaa Udeshi, Shelby Smout, Madison Caballero, Amy Rapp, Alexander Kolevzon, Behrang Mahjani, Seulgi Jung

## Abstract

Rare deleterious variants in *SHANK3* are established causes of autism spectrum disorder (ASD), but the extent to which they define a phenotypically and genetically coherent ASD subgroup remains unclear. Using the SPARK cohort, we identified 132 *SHANK3* variant carriers; 108 had phenotype data and were compared with 47,555 non-carrier ASD cases. *SHANK3* damaging variant carriers showed lower cognitive ability, poorer motor coordination, and delayed developmental milestones. Protein-truncating variant and deletion carriers showed similarly severe phenotypic profiles, whereas duplication carriers did not differ from non-carriers. A combined threshold of intelligence quotient (IQ) < 70 and impaired motor coordination (DCDQ total score) < 35 defined a discriminative cognitive-motor phenotype among cases meeting this cognitive-motor phenotype. Beyond *SHANK3*, *SLC6A1* was the only additional gene reaching false discovery rate significance, while pathway analyses implicated synaptic and chromatin-related processes. Phenotype-meeting cases did not show elevated ASD polygenic risk, supporting a rare-variant-enriched cognitive-motor subgroup within ASD.

## Introduction

Autism spectrum disorder (ASD) is characterized by marked phenotypic heterogeneity, with substantial variability in cognitive ability, developmental trajectories, and associated neurobehavioral features across affected individuals^1–3^. This clinical diversity is mirrored by a complex genetic architecture in which rare variants of large effect and common polygenic variation both contribute to liability^4,5^. A central challenge in ASD research is to determine whether specific genetic etiologies define more phenotypically and biologically coherent subtypes within this heterogeneous disorder.

Among rare variants implicated in ASD, loss-of-function (LoF) variants in *SHANK3* (SH3 and multiple ankyrin repeat domains 3) are among the most well-established genetic causes^6,7^. *SHANK3* encodes a synaptic scaffolding protein that is essential for postsynaptic density organization at glutamatergic synapses.^8,9^ Through its multiple protein interaction domains, *SHANK3* links glutamate receptors to intracellular signaling cascades and the actin cytoskeleton, and its disruption impairs synaptic plasticity and dendritic spine morphology^8^. Haploinsufficiency of *SHANK3*, either through protein-truncating or through 22q13.3 deletions (Phelan-McDermid syndrome), is associated with a characteristic clinical profile that includes intellectual disability, neonatal hypotonia, motor impairment, and autism-related features^10–12^. Cognitive impairment is highly prevalent, with the majority of affected individuals showing moderate to severe intellectual disability^10,13^, and motor deficits including neonatal hypotonia reported in over 97% of cases^12^. Because *SHANK3* occupies a central position in the postsynaptic scaffold, it provides a particularly informative model for linking genetic variation to clinical presentation and synaptic biology in ASD.

Despite the well-described clinical consequences of *SHANK3* haploinsufficiency, important gaps remain. The specific phenotypic dimensions that most clearly distinguish *SHANK3* carriers from the broader ASD population have not been systematically evaluated using the same standardized assessments. Previous phenotypic studies have largely focused on clinically ascertained cohorts with Phelan-McDermid syndrome^10,14,15^, and systematic comparisons of *SHANK3* carriers with other individuals with ASD remain limited. In addition, it remains unclear whether ASD individuals who share a *SHANK3*-like phenotypic profile, despite lacking *SHANK3* variants, also show genetic convergence in related biological pathways. Finally, the extent to which common polygenic variation contributes to ASD liability in individuals with *SHANK3*-associated phenotypes, as compared to the broader ASD population, has not been examined.

Here, we used the SPARK cohort to systematically characterize the phenotypic profile of ASD cases carrying rare deleterious *SHANK3* variants, identify discriminative phenotypic thresholds, test whether ASD cases meeting this phenotypic profile are enriched for rare deleterious variants in related genes, and evaluate the contribution of common polygenic risk across ASD subtypes defined by this profile.

## Results

### *SHANK3* Variant Carriers in SPARK

From the SPARK cohort, we identified 132 ASD cases carrying 133 rare deleterious *SHANK3* variant occurrences, including 34 protein-truncating variant (PTV) occurrences in 34 carriers, 56 deletions in 56 carriers, 7 pathogenic missense variant occurrences in 6 carriers, and 36 duplications in 36 carriers (**Table S1**). Thus, the number of variant occurrences exceeded the number of carriers by one because one individual carried two pathogenic missense variants. Of the 56 deletion carriers, 10 overlapped with *SHANK3* gCNV cases previously reported in Fu et al., 2022^16^. The remaining 53,289 ASD cases without *SHANK3* variants served as the comparison group.

Among 40 individuals carrying PTVs or pathogenic missense variants, we observed 41 variant occurrences, corresponding to 32 unique sequence variants: 14 frameshift, 8 nonsense, 5 splice-site, and 5 missense variants (**Fig. 1A**). These variants were distributed across the full length of the *SHANK3* protein, with recurrent variants observed at several positions: the missense variant R753H and the frameshift variant p.Leu1285fs in the proline-rich region each occurred in three independent individuals.

**Fig. 1:**
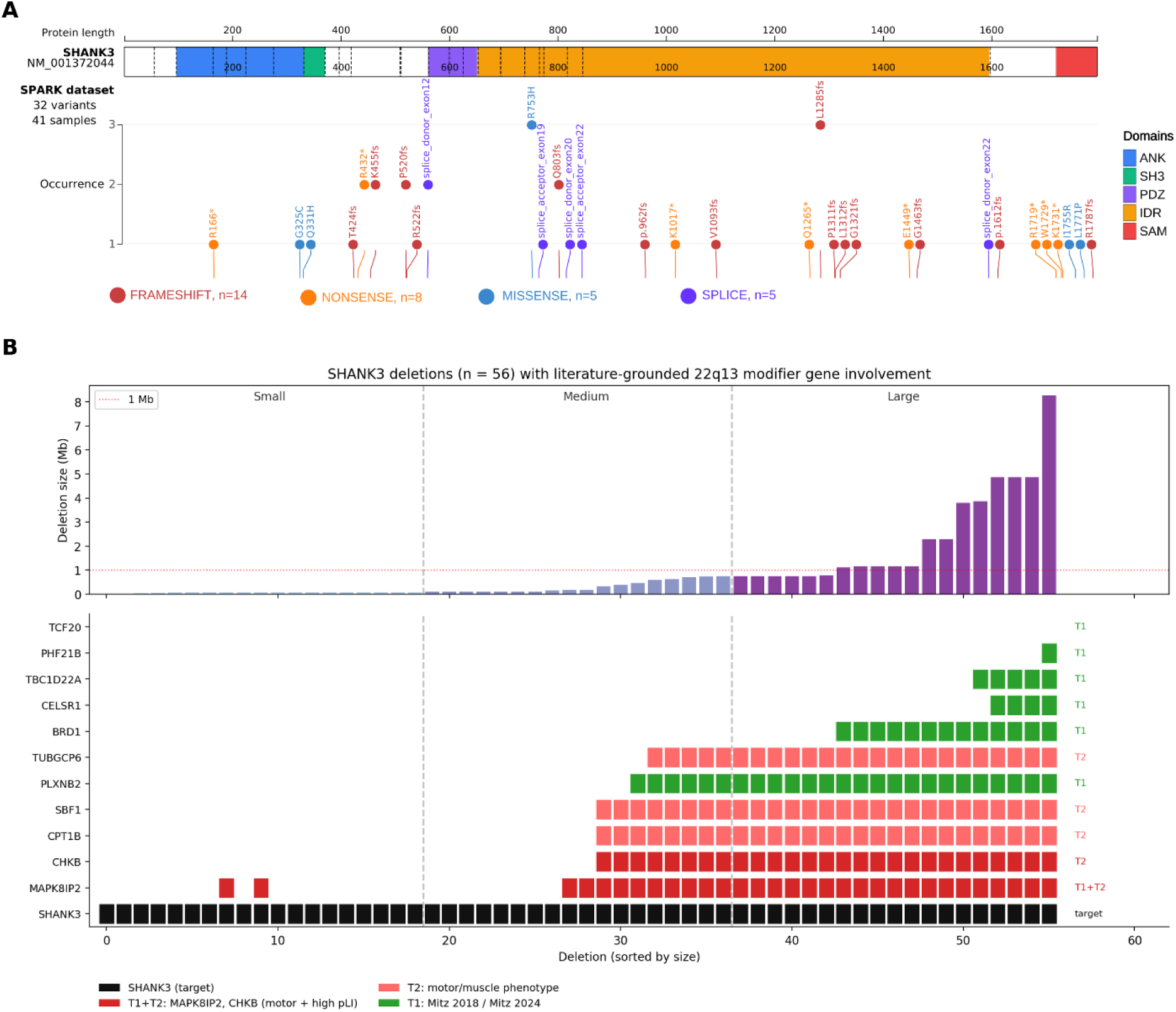
Landscape of rare deleterious *SHANK3* variants identified in ASD cases from the SPARK cohort. **(A)** Protein-truncating and missense variants plotted along the *SHANK3* protein (NM_001372044). Functional domains: ankyrin repeats (ANK, blue), Src homology 3 (SH3, green), PSD-95/Dlg/ZO-1 (PDZ, purple), intrinsically disordered region (IDR, orange), and sterile alpha motif (SAM, red). The y-axis indicates the number of independent occurrences of each variant. A total of 32 unique variants were identified across 41 samples, comprising 14 frameshift, 8 nonsense, 5 missense, and 5 splice-site variants. Plot generated using ProteinPaint (https://proteinpaint.stjude.org/). **(B)** Size distribution of 56 *SHANK3*-overlapping deletions and involvement of literature-grounded 22q13.3 modifier genes. Top: deletion sizes sorted from smallest to largest, colored by size tertile (Small < 0.10 Mb, Medium 0.10–0.75 Mb, Large > 0.75 Mb). Bottom: raster plot showing overlap of each deletion with *SHANK3* (target, black) and 11 candidate PMS modifier genes prioritized by Mitz et al., 2018 and Mitz et al., 2024.^17,18^ Tier 1 (T1, green): high-confidence neurodevelopmental modifiers (pLI > 0.9, PIF > 0.5); Tier 2 (T2, salmon): motor/muscle phenotype relevance; Tier 1+2 overlap (red): *MAPK8IP2*, whose murine knockout shows cerebellar motor deficits,^30^ and *CHKB*, whose biallelic loss causes congenital muscular dystrophy.^31^

Among the 56 deletion carriers, deletion sizes spanned three orders of magnitude, from 10 kb to 8.3 Mb (median 0.18 Mb, mean 0.94 Mb), with no preferred breakpoint (**Fig. 1B, Table S2**). To assess the extent to which these deletions extended beyond *SHANK3* into adjacent 22q13.3 genes with potential phenotypic relevance, we annotated each deletion against a literature-curated set of 22q13.3 modifier genes prioritized by Mitz et al. on the basis of pLI scores, population impact factor (PIF), and functional evidence in PMS (**Table S3**).^17,18^ Of the 56 deletions, 25 (45%) were *SHANK3*-restricted, with no overlap of any prioritized neighboring modifier gene. The remaining 31 (55%) extended into one or more candidate modifiers, most commonly *MAPK8IP2* (n = 31, 55%), *CHKB* (n = 27, 48%), *SBF1* (n = 27, 48%), *CPT1B* (n = 27, 48%), *PLXNB2* (n = 25, 45%), and *TUBGCP6* (n = 24, 43%). Deletions in the upper size tertile (> 0.75 Mb, n = 19) uniformly disrupted all of these neighboring genes, whereas deletions in the lower tertile (< 0.10 Mb, n = 19) were largely *SHANK3*-restricted (**Fig. 1B, Table S4**).

Of the 132 *SHANK3* variant carriers, 108 had at least one phenotypic measure available (54 deletion, 21 duplication, 5 missense, and 28 PTV) and were compared against 47,555 non-carrier ASD cases. The heatmap of effect sizes across variant types (**Figure 2 and Table S5**) illustrated a clear distinction between variant categories. Deletion and PTV carriers displayed consistently strong negative effect sizes (blue) across motor coordination and cognitive variables, and positive effect sizes (red) across developmental milestone delays, with the majority reaching Benjamini-Hochberg (BH)-adjusted significance. Missense carriers showed a similar directional pattern to deletion and PTV carriers, particularly in motor domains, though few results reached statistical significance given the small sample size (n = 5). In contrast, duplication carriers showed uniformly weak and non-significant effect sizes across all phenotypic variables, with no variable reaching even nominal significance. Importantly, direct pairwise comparisons between PTV and deletion carriers revealed no significant differences in any phenotypic variable after multiple testing correction (all BH-adjusted *P* > 0.05; **Table S5**), suggesting that the observed phenotype is consistent across structurally distinct *SHANK3* loss-of-function variant classes regardless of deletion extent, although power to detect modifier effects was limited. Given that duplication carriers did not exhibit the phenotypic profile characteristic of *SHANK3* damaging variants, subsequent analyses focused on PTV, deletion, and missense.

**Figure 2.**
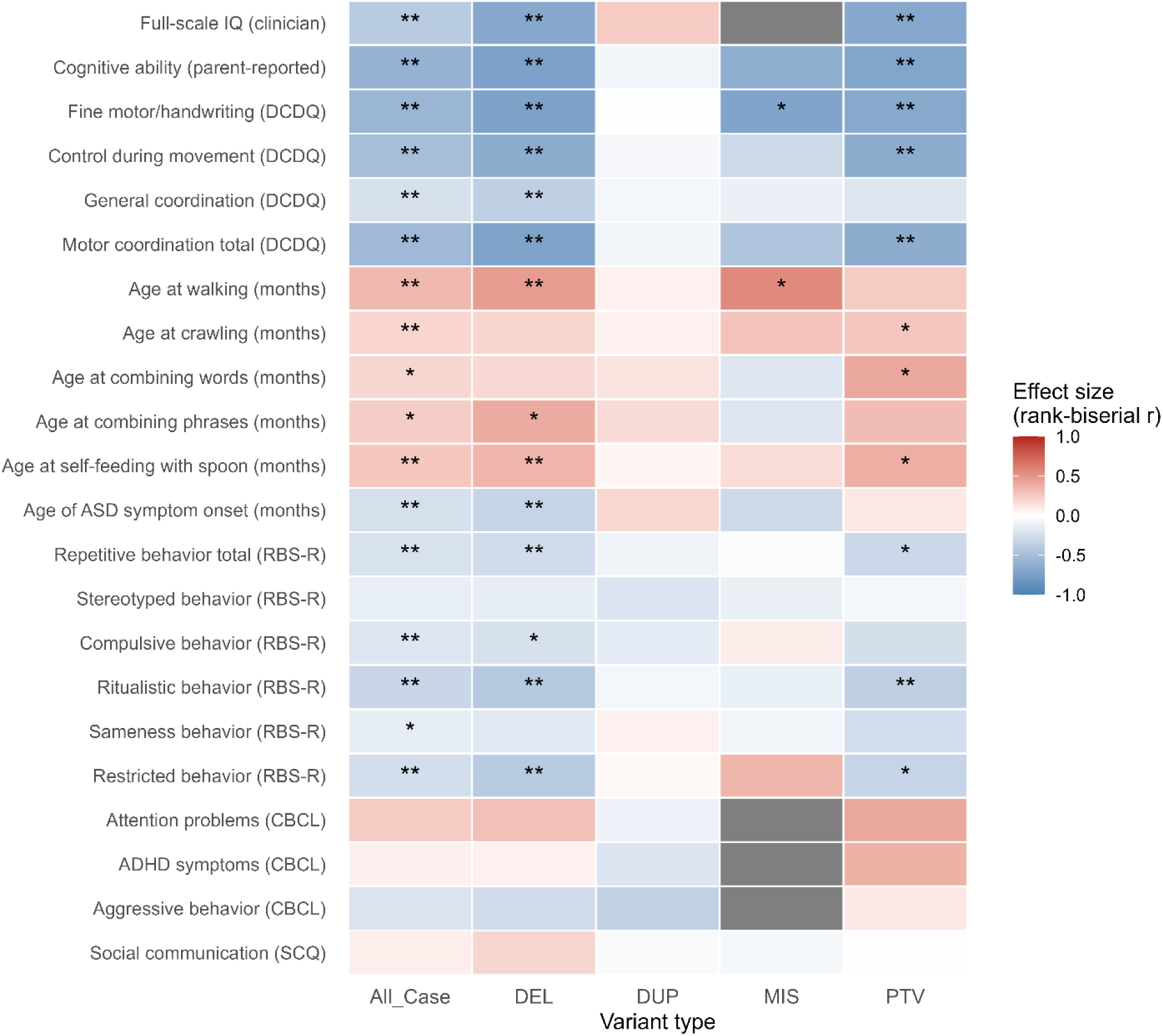
Effect sizes of phenotypic comparisons between *SHANK3* variant carriers and non-carrier ASD cases. Heatmap displaying rank-biserial correlation coefficients (r) from Wilcoxon rank-sum tests for each phenotypic variable (rows) across variant types (columns): all carriers combined (All_Case), deletion (DEL), duplication (DUP), missense (MIS), and protein-truncating variant (PTV) carriers, each compared against non-carrier ASD cases. Blue indicates lower scores or earlier ages in carriers relative to non-carrier ASD cases; red indicates higher scores or later ages in carriers. Grey cells indicate comparisons where insufficient data were available. Asterisks denote statistical significance (*nominal *P* < 0.05; **Benjamini-Hochberg adjusted *P* < 0.05).

When *SHANK3* damaging variant carriers (PTV, deletion, and missense combined) were compared against non-carrier ASD cases, significant differences were observed across multiple phenotypic domains (**Figure 3 and Table S6**). All *P* values reported below are BH-adjusted unless otherwise noted. The largest effect sizes were in cognitive ability, with parent-reported cognitive scores substantially lower in *SHANK3* damaging variant carriers (median 47 vs. 84.5, r = −0.72, *P* = 1.7 × 10^-9^), and fine motor/handwriting (median 4 vs. 9, r = −0.71, *P* = 5.1 × 10^-13^). Clinician-administered FSIQ was also significantly lower (median 49 vs. 81, r = −0.68, *P* = 0.001). DCDQ total score, which combines fine motor, control during movement, and general coordination subscales into a single measure of overall motor coordination (median 24 vs. 36, r = −0.66, *P* = 2.2 × 10^-11^), control during movement (median 10 vs. 16, r = −0.62, *P* = 2.9 × 10^-11^), and general coordination (median 9 vs. 11, r = −0.28, *P* = 0.007) were all significantly impaired. Developmental milestones were significantly delayed, including walking age (median 15.5 vs. 13 months, r = 0.40, *P* = 1.4 × 10^-6^), self-feeding with a spoon (median 24 vs. 16 months, r = 0.34, *P* = 6.4 × 10^-4^), and crawling age (median 9 vs. 8 months, r = 0.23, *P* = 0.009). *SHANK3* damaging variant carriers also exhibited lower scores on multiple RBS-R subscales relative to non-carrier ASD cases, including ritualistic behavior (r = −0.38, *P* = 4.4 × 10^-6^), restricted behavior (r = −0.31, *P* = 1.8 × 10^-4^), RBS-R total score (r = −0.26, *P* = 0.002), compulsive behavior (r = −0.21, *P* = 0.016), and sameness behavior (r = −0.19, *P* = 0.025), with lower scores indicating fewer parent-reported repetitive behaviors despite greater overall clinical severity. Age of ASD symptom onset was earlier in *SHANK3* damaging variant carriers (median 14 vs. 18 months, r = −0.25, *P* = 0.003). CBCL attention problems were elevated in *SHANK3* damaging variant carriers (r = 0.33, *P* = 0.045). In total, 16 phenotypic variables reached BH-adjusted significance at *P* < 0.05.

**Figure 3.**
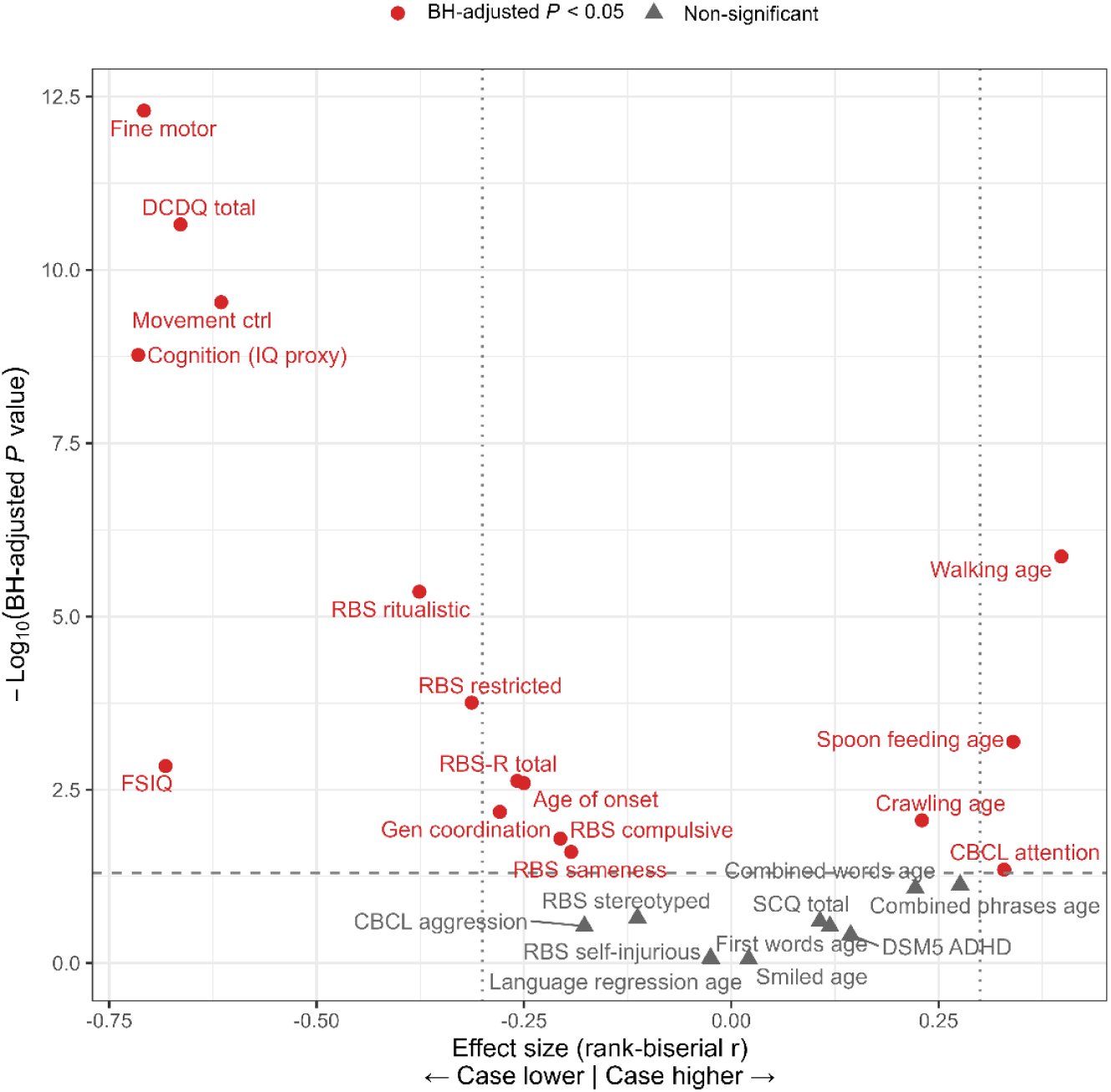
Volcano plot of phenotypic differences between *SHANK3* damaging variant carriers and non-carrier ASD cases. Each point represents a phenotypic variable compared between *SHANK3* damaging variants (PTV, deletion, and missense combined) and non-carrier ASD cases using Wilcoxon rank-sum tests. The x-axis shows the effect size (rank-biserial r), where negative values indicate lower scores in *SHANK3* damaging variant carriers and positive values indicate higher scores or later developmental milestone ages. The y-axis shows the negative log_10_-transformed Benjamini-Hochberg adjusted *P* value. Red circles denote variables reaching BH-adjusted significance (*P* < 0.05); grey triangles denote non-significant variables. The horizontal dashed line indicates the BH-adjusted *P* = 0.05 threshold. Vertical dotted lines mark effect sizes of r = -0.3 and r = 0.3. Motor coordination (fine motor, DCDQ total, control during movement) and cognitive ability showed the largest effect sizes and strongest statistical significance in the upper left quadrant, while delayed developmental milestones (walking age, spoon feeding age) appeared in the upper right.

### Phenotypic Threshold Sensitivity Analysis

To explore whether combinations of phenotypic measures could discriminate *SHANK3* damaging variant carriers from non-carrier ASD cases, we performed a systematic two-measure threshold sweep across 16 phenotypic variables that showed significant associations with *SHANK3* damaging variant status. Individual ROC analysis revealed that cognitive ability (AUC = 0.858), fine motor/handwriting (AUC = 0.854), FSIQ (AUC = 0.841), and DCDQ total score (AUC = 0.832) were the strongest individual discriminators (**Table S7**).

Among all pairwise threshold combinations, the top-performing pairs consistently involved cognitive ability combined with motor coordination measures (**Table S8**). Given that the DCDQ total score captures motor impairment more comprehensively than a single subscale and is more widely used in clinical settings, and that cognitive impairment and motor deficits are the two most consistently reported phenotypes in *SHANK3*-related disorders,^10,13,19,20^ we selected cognitive ability and DCDQ total score as the two-measure combination for further evaluation. A grid search across IQ and DCDQ thresholds (**Table S9**) identified IQ < 70 and DCDQ < 35 as providing a favorable balance of sensitivity and specificity.

Among the 13 *SHANK3* damaging variant carriers with complete data on both measures, 12 (92.3%) met the combined criteria, compared to 15.2% of non-carrier ASD cases (1,409 of 9,279), corresponding to an odds ratio of 67.0 (95% CI: 9.9 to 2,819.7, Fisher’s exact *P* = 1.8 x 10^-9^). ROC analysis comparing individual and combined measures suggested that the combination of IQ and DCDQ (AUC = 0.903) may improve discrimination over either measure alone (IQ: AUC = 0.884; DCDQ: AUC = 0.860). However, these results should be interpreted with caution given the small number of *SHANK3* damaging variant carriers with complete data on both measures (n = 13), as reflected by the wide confidence interval of the odds ratio. Validation in larger, independent cohorts will be necessary to confirm the clinical utility of this phenotypic combination as a screening tool for *SHANK3* phenotype ASD.

### Gene-level Enrichment in *SHANK3* phenotype Cases

Among ASD cases with complete data on both cognitive ability and DCDQ total score, 1,421 met the *SHANK3* phenotype criteria (IQ < 70 and DCDQ < 35) and 7,871 did not. Gene-level enrichment analysis was performed comparing carriers of rare PTVs or deletions between the two groups (without pathogenic missense variants or duplications), restricted to 255 ASD risk genes with FDR < 0.1 from the TADA analysis in the Fu et al., 2022 (**Table 1 and Table S10**)^16^. *SHANK3* was the most significantly enriched gene (10 carriers vs. 1, FDR = 5.7 x 10^-6^), which is expected given that the phenotypic criteria were informed by *SHANK3* carrier characteristics and thus serves as positive control rather than an independent finding. More notably, *SLC6A1* was the only additional gene reaching FDR < 0.05 (4 carriers vs. 0, FDR = 0.024), encoding the GABA transporter GAT-1, a key mediator of synaptic GABA reuptake, with pathogenic variants known to cause neurodevelopmental disorders including epilepsy, intellectual disability, and autism-related features^21–23^. *SETD1B,* which encodes a lysine-specific histone H3K4 methyltransferase and has been implicated in a neurodevelopmental disorder characterized by intellectual disability, language delay, and epilepsy^24,25^, showed a trend toward enrichment (3 vs. 0, FDR = 0.106). Additional nominally significant genes (*P* < 0.05) included *DLG4*, *EHMT1*, *TCF4*, *NOTCH1*, *ADCY5*, and *AUTS2*.

**Table 1.** Gene-level enrichment of rare deleterious variants (protein-truncating variants or deletions) in ASD risk genes among *SHANK3*-phenotype cases (IQ < 70 and DCDQ < 35; n = 1,421) compared with non-phenotype ASD cases (n = 7,871).

| Gene | *SHANK3*-phenotype |  | Non-phenotype |  | OR | *P* value | FDR |
| --- | --- | --- | --- | --- | --- | --- | --- |
|  | *n* | (%) | *n* | (%) |  |  |  |
| *SHANK3* | 10 | 0.70 | 1 | 0.01 | 55.75 | $6.46 \times 10^{-8}$ | $5.7 \times 10^{-6}$ |
| *SLC6A1* | 4 | 0.28 | 0 | 0.00 | - | $5.45 \times 10^{-4}$ | 0.024 |
| *SETD1B* | 3 | 0.21 | 0 | 0.00 | - | $3.57 \times 10^{-3}$ | 0.106 |
| *DLG4* | 2 | 0.14 | 0 | 0.00 | - | $2.34 \times 10^{-2}$ | 0.260 |
| *EHMT1* | 2 | 0.14 | 0 | 0.00 | - | $2.34 \times 10^{-2}$ | 0.260 |
| *TCF4* | 2 | 0.14 | 0 | 0.00 | - | $2.34 \times 10^{-2}$ | 0.260 |
| *NOTCH1* | 2 | 0.14 | 0 | 0.00 | - | $2.34 \times 10^{-2}$ | 0.260 |
| *ADCY5* | 2 | 0.14 | 0 | 0.00 | - | $2.34 \times 10^{-2}$ | 0.260 |
| *AUTS2* | 3 | 0.21 | 2 | 0.03 | 8.32 | $2.80 \times 10^{-2}$ | 0.277 |

Genes with nominal $P < 0.05$ are shown, ordered by $P$ value. Carrier rates are expressed as percentages. OR, odds ratio from Fisher's exact test; FDR, Benjamini–Hochberg false discovery rate. ∞ indicates that no carriers were observed in the non-phenotype group.

### Pathway Analysis

To explore the biological processes represented by the nine nominally significant genes (*P* < 0.05 from the gene-level enrichment analysis), we performed pathway enrichment analysis across four databases (**Figure 4 and Table S11**). All *P* values reported below are Benjamini-Hochberg adjusted.

**Figure 4.**
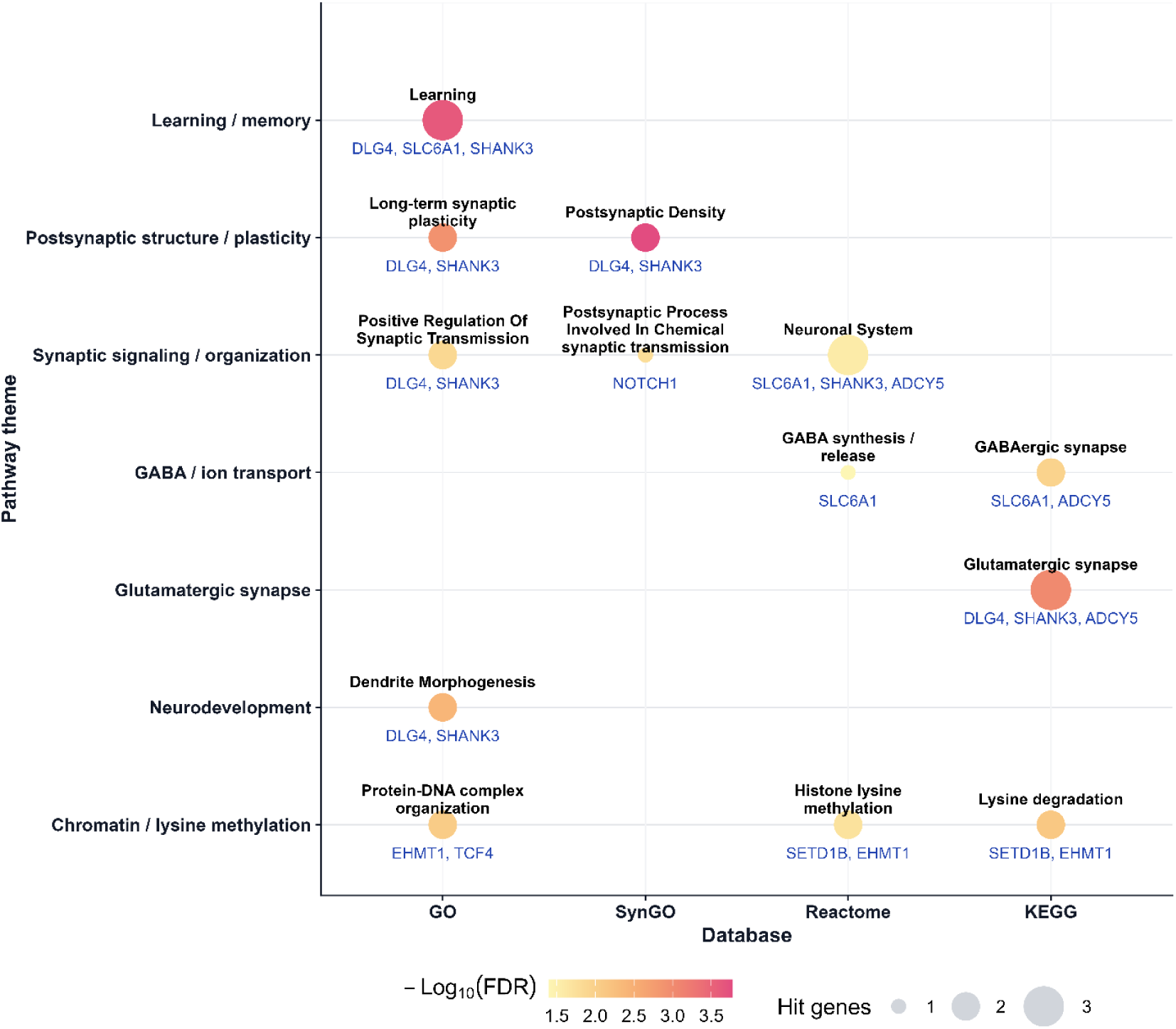
Convergent pathway enrichment of *SHANK3* phenotype-associated genes. Representative pathway-enrichment themes across GO, SynGO, Reactome, and KEGG for *SHANK3* phenotype-associated genes. Each circle denotes the top representative term for a given theme and database. Circle size indicates the number of overlapping genes contributing to the enrichment, and circle color indicates enrichment significance as -log_10_(FDR). Bold text above each circle shows the representative term, and blue text below indicates the overlapping genes. Enrichment signals converge on learning/memory, postsynaptic structure and plasticity, synaptic signaling and organization, presynaptic release/vesicle pathways, GABA/ion transport, glutamatergic synapse, neurodevelopment, and chromatin/lysine methylation.

In the SynGO database^26^, the most significantly enriched term was structural constituent of postsynaptic density (*DLG4*, *SHANK3*; *P* = 1.7 x 10^-4^), followed by postsynaptic process involved in chemical synaptic transmission (*NOTCH1*; *P* = 0.023) and regulation of postsynapse organization (*SHANK3*; *P* = 0.037). GO Biological Process^27^ analysis identified learning (*DLG4*, *SLC6A1*, *SHANK3*; *P* = 2.4 x 10^-4^), regulation of long-term neuronal synaptic plasticity (*DLG4*, *SHANK3*; *P* = 8.6 x 10^-4^), and dendritic spine morphogenesis (*DLG4*, *SHANK3*; *P* = 1.5 x 10^-3^) as top enriched terms, alongside protein-DNA complex organization (*EHMT1*, *TCF4*; *P* = 0.011). In the Kyoto Encyclopedia of Genes and Genomes (KEGG) database,^28^ glutamatergic synapse was the most enriched pathway (*DLG4*, *SHANK3*, *ADCY5*; *P* = 9.5 x 10^-4^), followed by lysine degradation (*SETD1B*, *EHMT1*; *P* = 5.3 x 10^-3^) and GABAergic synapse (*SLC6A1*, *ADCY5*; *P* = 0.011). Reactome^29^ analysis highlighted histone lysine methylation (*SETD1B*, *EHMT1*; *P* = 0.019) and neuronal system (*SLC6A1*, *SHANK3*, *ADCY5*; *P* = 0.023).

Collectively, these results indicate that ASD cases meeting the *SHANK3*-associated phenotypic profile are enriched for rare deleterious variants in genes that converge on postsynaptic density organization, glutamatergic synaptic transmission, and chromatin remodeling.

### Polygenic Risk Score Analysis

To examine whether common variant burden differs between *SHANK3* phenotype cases (IQ < 70 and DCDQ < 35) and non-*SHANK3* phenotype cases, we compared polygenic risk scores (PRS) for ASD, ADHD, OCD, and schizophrenia across three groups. After restricting to individuals of non-Finnish European (NFE) ancestry, 753 *SHANK3* phenotype ASD cases, 5,192 non-*SHANK3* phenotype ASD cases, and 8,198 unaffected controls were included in the analysis (**Figure 5 and Table S12**).

**Figure 5.**
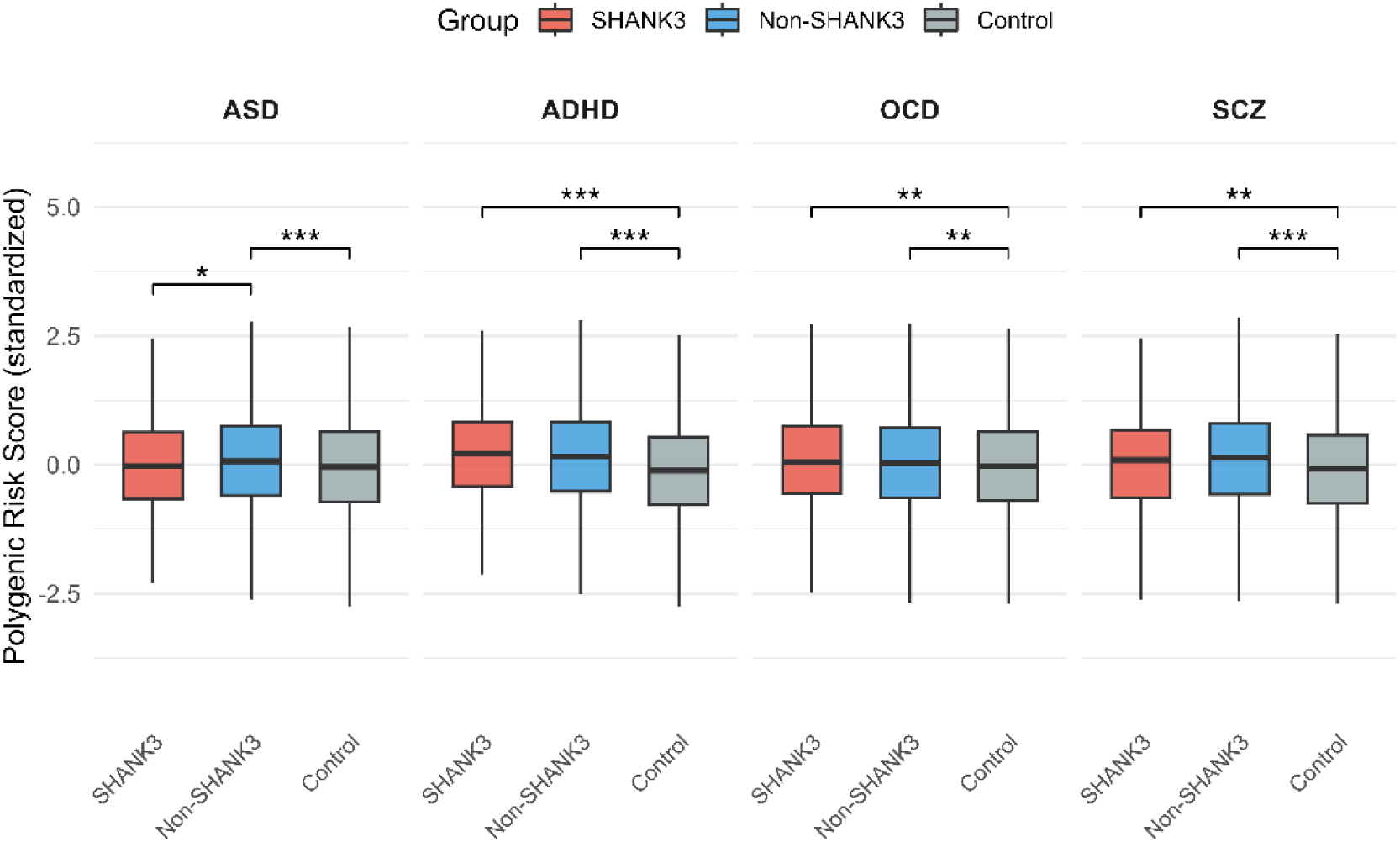
Polygenic risk score comparisons across *SHANK3* phenotype ASD cases, non-*SHANK3* phenotype ASD cases, and unaffected controls. Boxplots show standardized polygenic risk scores (PRS) for autism spectrum disorder (ASD), attention-deficit/hyperactivity disorder (ADHD), obsessive-compulsive disorder (OCD), and schizophrenia (SCZ) across three groups of European ancestry: *SHANK3* phenotype ASD cases (red, n = 753), non-*SHANK3* phenotype ASD cases (blue, n = 5,192), and unaffected controls (grey, n = 8,198). Pairwise comparisons were performed using Wilcoxon rank-sum tests with Benjamini-Hochberg correction within each PRS type. Only significant comparisons are shown (*BH-adjusted *P* < 0.05; **BH-adjusted *P* < 0.01; ***BH-adjusted *P* < 0.001).

For ASD PRS, non-*SHANK3* phenotype ASD cases showed significantly higher scores compared to controls (*P* = 2.8 x 10^-10^), consistent with a polygenic contribution to their ASD liability. In contrast, *SHANK3* phenotype cases did not differ from controls in ASD PRS (*P* = 0.583) and had significantly lower ASD PRS than non-*SHANK3* phenotype ASD cases (*P* = 0.018). This pattern suggests that ASD in *SHANK3* phenotype cases could be primarily driven by rare variant effects rather than common polygenic risk.

For ADHD PRS, both ASD groups showed significantly elevated scores compared to controls (*SHANK3* phenotype: *P* = 1.3 x 10^-16^; non-*SHANK3* phenotype: *P* = 1.5 x 10^-52^), with no significant difference between the two ASD groups (*P* = 0.342). Similarly, OCD and schizophrenia PRS were elevated in both ASD groups relative to controls (OCD: non-*SHANK3* phenotype *P* = 6.0 × 10^-3^, *SHANK3* phenotype *P* = 7.5 × 10^-3^; schizophrenia: non-*SHANK3* phenotype *P* = 6.0 × 10^-28^, *SHANK3* phenotype *P* = 1.5 × 10^-3^), with no significant differences between the two ASD groups (OCD *P* = 0.189; schizophrenia *P* = 0.054). The elevation of ADHD, OCD, and schizophrenia PRS in both ASD groups relative to controls, with no significant differences between the two ASD groups, suggests a shared cross-disorder polygenic burden regardless of *SHANK3* variant status.

## Discussion

In this study, we show that rare damaging *SHANK3* variants are associated with a distinctive phenotypic profile within ASD, characterized primarily by cognitive impairment and motor dysfunction. In the SPARK cohort, *SHANK3* damaging variant carriers exhibited substantially lower cognitive ability, poorer motor coordination, and delayed developmental milestones compared with non-carrier ASD cases. Beyond describing the phenotype of *SHANK3* carriers, our results further suggest that this clinical profile captures a biologically informative subgroup of ASD: individuals meeting *SHANK3*-associated phenotypic criteria were enriched for rare deleterious variants in genes converging on synaptic and chromatin-related pathways.

The phenotypic comparisons identified cognitive impairment and motor dysfunction as the clearest features distinguishing *SHANK3* damaging variant carriers from the broader ASD population, consistent with the well-established clinical presentation of Phelan-McDermid syndrome^10,12,19^. Deletion and PTV carriers showed the broadest and most severe impairments, whereas duplication carriers did not differ significantly from non-carrier ASD cases. This contrast supports the interpretation that the observed phenotype is primarily related to loss-of-function rather than to altered *SHANK3* dosage more generally, at least within this dataset.

*SHANK3* variant carriers in this cohort included 54 deletions ranging from 10 kb to 8.3 Mb, with larger deletions extending into multiple 22q13.3 genes prioritized as candidate PMS modifiers (**Fig. 1B, Tables S2-S4**).^17,18^ Deletions above 0.75 Mb uniformly involved *MAPK8IP2*, *CHKB*, *PLXNB2*, and several additional candidate modifiers. This raises the possibility that phenotype contributions from adjacent genes such as *MAPK8IP2*, whose knockout in mice produces motor and cerebellar deficits,^30^ and *CHKB*, whose biallelic loss causes megaconial congenital muscular dystrophy in humans,^31^ could amplify the observed signature in larger-deletion carriers. However, direct pairwise comparisons between PTV carriers, which disrupt *SHANK3* specifically, and deletion carriers, which often co-delete neighboring genes, revealed no significant phenotypic differences after multiple testing correction (**Table S5**). This null finding supports the interpretation that *SHANK3* loss of function itself accounts for the bulk of the observed phenotype. Subtle modifier contributions cannot be fully excluded, but the overall signature appears robust to deletion extent.

Developmental milestones, including walking, crawling, and self-feeding, were also delayed in *SHANK3* damaging variant carriers, further supporting the view that *SHANK3*-related ASD is marked by broad neurodevelopmental impairment rather than isolated differences in core autism features. These findings are also biologically plausible given prior evidence linking *SHANK3* to neuromuscular junction maturation and skeletal muscle function, which may contribute to the prominence of motor deficits in affected individuals^20^.

*SHANK3* damaging variant carriers also had lower scores on multiple RBS-R subscales than non-carrier ASD cases. At first glance, this may appear counterintuitive, since *SHANK3*-related disorders are strongly associated with neurodevelopmental impairment and autism-related features. However, lower scores on subscales involving cognitively mediated repetitive behaviors, such as rituals, compulsions, and insistence on sameness, may reflect reduced expression or reduced detectability of these behaviors in individuals with more severe cognitive and motor impairment.^32–34^ In this context, the *SHANK3*-associated phenotype may be distinguished less by increased repetitive behavior burden overall than by a combination of intellectual disability, motor dysfunction, and delayed developmental acquisition.

A key finding of this study is that phenotype-based stratification identified a rare-variant-enriched ASD subgroup with convergent biological signal. Among ASD cases meeting the *SHANK3*-associated phenotype criteria, *SHANK3* itself remained the gene with the largest absolute number of variant carriers (n = 12), followed by *SLC6A1* (n = 4) as the only additional gene reaching FDR significance. However, even together these two genes accounted for only a small fraction of phenotype-meeting cases (< 1.5%), indicating that the severe cognitive-motor phenotype is characteristic of *SHANK3* loss of function but not specific to it. Similar clinical presentations likely arise through rare-variant disruption of a broader set of neurodevelopmental disorder (NDD)-associated genes, consistent with the nominal enrichment of *SETD1B*, *DLG4*, *NOTCH1*, *EHMT1*, and *TCF4* in this phenotype group. *SLC6A1* encodes the GABA transporter GAT-1, which regulates synaptic GABA reuptake and inhibitory neurotransmission,^21,22^ its enrichment alongside *SHANK3* suggests that the severe cognitive-motor phenotype may reflect convergent disruption of both excitatory and inhibitory synaptic function.

Pathway analyses across SynGO,^26^ GO,^27^ Reactome,^29^ and KEGG^28^ implicated postsynaptic density organization, glutamatergic synaptic signaling, GABAergic neurotransmission, and chromatin remodeling. Sensitivity analysis excluding *SHANK3* itself from the enriched gene set (**Table S13**) showed that the core pathway signals were largely retained: GABAergic synapse (driven by *SLC6A1* and *ADCY5*), glutamatergic synapse (*DLG4* and *ADCY5*), and chromatin-related pathways (*SETD1B*, *EHMT1*, *TCF4*) remained significant at BH-adjusted *P* < 0.05, indicating that convergence on synaptic and chromatin biology is not solely driven by *SHANK3* itself. Postsynaptic density signals attenuated but persisted through *DLG4*, consistent with partial but not complete *SHANK3*-dependence of this particular signal. These results support a biologically meaningful convergence across rare-variant-enriched genes in the severe cognitive-motor phenotype group.

The PRS analyses provided complementary insight into the genetic architecture of this phenotype-defined subgroup. Non-*SHANK3* phenotype ASD cases showed significantly elevated ASD PRS relative to controls, whereas *SHANK3* phenotype cases did not. Although this result should be interpreted cautiously, it is consistent with the possibility that ASD liability in *SHANK3* phenotype cases is less strongly indexed by current ASD polygenic risk scores^4,35^ and may be more strongly influenced by rare variants of larger effect. By contrast, both ASD groups showed elevated PRS for ADHD, OCD, and schizophrenia relative to controls, with no significant differences between the two ASD subgroups. This pattern suggests that cross-disorder common variant liability may be broadly shared across ASD subtypes, even when the contribution of ASD-specific polygenic burden differs. This observation is consistent with prior evidence of substantial genetic overlap between ASD and other neurodevelopmental and psychiatric conditions,^35^ and suggests that shared risk for co-occurring conditions such as ADHD and OCD may characterize *SHANK3*-associated ASD as well as broader ASD populations. Clinically, this implies that individuals with a *SHANK3*-associated phenotype may carry comparable polygenic risk for comorbid neuropsychiatric conditions, which warrants consideration in long-term clinical monitoring.

Several limitations should be considered. First, the number of *SHANK3* damaging variant carriers with complete data on both IQ and DCDQ was small, resulting in wide confidence intervals around the estimated effect sizes and limiting the stability of the threshold-based phenotype definition. Second, the key phenotype analyses relied on complete-case subsets, and missingness patterns were not formally evaluated; therefore, the discriminative performance of the selected phenotypic combination may have been influenced by ascertainment or non-random missingness patterns. Third, PRS analyses were restricted to individuals of non-Finnish European ancestry to minimize population stratification, which limits generalizability across ancestries. Fourth, ASD diagnoses in SPARK were based on caregiver report, which may introduce phenotypic heterogeneity. Finally, the cross-sectional structure of the available data precluded analysis of developmental trajectories over time.

Building on prior characterizations of *SHANK3*-related phenotypes in clinically ascertained Phelan-McDermid syndrome cohorts,^10,14,15^ our study advances this work in three important ways. First, by leveraging the large SPARK research cohort, we systematically characterize the *SHANK3* phenotype relative to the broader ASD population rather than the general population, thereby identifying the specific phenotypic dimensions that distinguish *SHANK3* carriers from other individuals with ASD. Second, we show that phenotype-guided stratification, without reference to genotype, recovers biologically coherent genetic convergence, including the identification of *SLC6A1* as enriched among ASD cases sharing the *SHANK3*-associated profile. Third, we demonstrate a dissociation between rare variant enrichment and common polygenic burden: ASD cases meeting the *SHANK3* phenotype show strong rare variant signal but attenuated ASD PRS, supporting a primarily rare-variant-driven architecture for this subtype. Together, these findings support a model in which rare deleterious *SHANK3* variants delineate a clinically recognizable, rare-variant-enriched ASD subtype centered on cognitive impairment and motor dysfunction, and they illustrate how phenotype-guided stratification may help parse ASD heterogeneity into biologically interpretable subgroups. More broadly, these findings provide a phenotype-guided framework that may inform genetic testing prioritization in clinical settings and help identify rare-variant-enriched subgroups that could benefit from mechanism-based intervention strategies. Replication in larger, independent cohorts and across diverse ancestries will be needed to confirm the generalizability of this phenotypic profile and its utility for identifying ASD cases with convergent synaptic pathway disruption.

## Methods

### Study Population

SPARK (Simons Foundation Powering Autism Research for Knowledge) is a large-scale cohort that compiles genetic, behavioral, and medical data from individuals with ASD and their families to advance understanding of autism spectrum disorder^36^. The third data release (July 2025), which incorporated whole exome sequencing (WES) and genome-wide association study (GWAS) data, contained 142,357 samples including 40,193 complete trios with 28,159 ASD-affected probands. All diagnoses in SPARK were based on caregiver report.

### Quality control of whole exome sequencing data

The most recent WES data (iWES v3), released in August 2024, comprised batches 1-9 ^36^. We performed comprehensive quality control (QC) on batches 1-4 and 5–9 at both genotype and sample levels, separately.

Genotype QC: We applied the following filters to exclude low-quality genotype calls: (1) read depth <10 or >1,000; (2) Y chromosome calls in female samples; (3) homozygous reference calls with <90% read depth supporting the reference allele or genotype quality (GQ) <25; and (4) homozygous alternate calls with <90% read depth supporting the alternate allele or PL[HomRef] <25. Heterozygous calls were removed if they met any of the following criteria: PL[HomRef] <25; call rate <90%; allele balance <25%; binomial probability of allele balance (centered on 0.5) <1×10⁻⁹; or location on the X or Y chromosome (excluding pseudoautosomal regions) in male samples.

Sample QC: We excluded samples with contamination rates >5%, call rates more than three standard deviations below the cohort mean, or duplicate status. Pedigree relationships were verified using KING-derived kinship coefficients (0.177–0.354 for first-degree relatives) ^37^. Incomplete trios were removed. For final variant QC, we excluded variants with call rate <90% or Hardy-Weinberg equilibrium *P* <10⁻¹². After QC, we retained 70,208 samples from batch 1-4 and 70,111 samples from batch 5-9.

Variant functional effects were annotated using the Variant Effect Predictor (VEP)^38^. Variants were categorized as protein-truncating variants (PTVs; frameshift, stop-gained, splice donor, and splice acceptor), missense, or synonymous. PTVs were retained only if designated "high-confidence" (HC) by the Loss-Of-Function Transcript Effect Estimator (LOFTEE)^39^, with all LOFTEE flags excluded except "SINGLE_EXON". Allele frequency was checked in both our case-control cohort and the non-psychiatric subset of gnomAD v3.1^39^.

### Quality control of copy-number variation data

Copy number variants (CNVs) were called from exome sequencing data of 142,128 individuals from the SPARK cohort (version 3) using XHMM (exome hidden Markov model)^40^. Due to the large sample size, CNV calling was performed across 10 batches.

Quality control procedures were applied to each batch following previously established protocols^41^. First, we retained only CNVs with quality scores ≥60, a threshold that has been shown to effectively filter out false-positive calls while maintaining sensitivity for true CNV detection. Second, to remove samples with an excess of CNV calls indicative of poor data quality or potential technical artifacts, we excluded individuals whose total CNV count exceeded three standard deviations above the batch-specific mean.

After applying these quality control filters, we retained 141,637 individuals (99.7% of the initial cohort). The pre-merge high-quality CNV call set comprised 84,043,360 CNVs, including 43,757,215 deletions and 40,286,145 duplications. Individual CNV calls were merged using a reciprocal 80% overlap criterion to consolidate redundant calls into unified CNV regions.

### Phenotypic Measures

Phenotypic data were obtained from SPARK^36^, including the Developmental Coordination Disorder Questionnaire (DCDQ)^42,43^, Repetitive Behavior Scale-Revised (RBS-R)^34^, Social Communication Questionnaire (SCQ)^44^, Child Behavior Checklist (CBCL)^45^, and Background History Questionnaire-Child/Dependent. The 26 instruments were selected based on their relevance to core ASD features and commonly reported phenotypes in *SHANK3*-related disorders.

The DCDQ assesses motor coordination across three subscales (control during movement, fine motor/handwriting, and general coordination), with lower scores indicating greater impairment; a total DCDQ score was also included. The RBS-R measures restricted and repetitive behaviors (RRBs) across six subscales (stereotyped, self-injurious, compulsive, ritualistic, sameness, and restricted behaviors) as well as a total score, with higher scores indicating greater severity. SCQ assesses social communication difficulties, with higher scores indicating greater impairment. From the CBCL, we extracted the Attention Problems, DSM-5 Attention-Deficit/Hyperactivity, and Aggressive Behavior scales; higher T-scores indicate greater symptom severity.

Cognitive ability was assessed using clinician-administered full-scale IQ (FSIQ) scores when available. For individuals without clinical IQ data, parent-reported cognitive test scores were used; these were recorded as categorical ranges (e.g., 70–79, 80–89), and midpoint values were assigned for analysis. Lower scores indicate greater cognitive impairment. Developmental milestones extracted from the Background History Questionnaire included ages at first smile, crawling, walking, self-feeding with a spoon, using single words, combining words, and combining phrases, with later ages indicating greater delay. Age of symptom onset and age of language regression were also extracted, with earlier regression indicating more severe disruption. Age at evaluation was recorded for each instrument to account for developmental differences across assessments.

### *SHANK3* Variants

To identify ASD cases with rare deleterious variants in *SHANK3*, we retained protein-truncating variants (PTVs), pathogenic missense variants (classified as "Pathogenic" or "Likely Pathogenic" in ClinVar) and CNVs with minor allele frequency < 0.0001 from both WES and CNV datasets. We identified a total of 132 ASD cases carrying 133 rare deleterious variants in *SHANK3*, including 34 PTVs, 7 pathogenic missense variants (in 6 carriers), 56 deletions, and 36 duplications. Of the 56 deletion carriers, 10 overlapped with *SHANK3* gCNV cases previously reported in Fu et al., 2022^16^. The remaining 53,289 ASD cases without *SHANK3* variants served as the comparison group. After restricting to individuals with at least one phenotypic measure available, 108 *SHANK3* variant carriers (54 deletion, 21 duplication, 5 missense, and 28 PTV) and 47,555 non-carrier ASD cases were retained for phenotypic analyses.

For each of the 56 *SHANK3*-overlapping deletions, deletion size was computed from breakpoint coordinates (GRCh38). Each deletion was annotated against a literature-curated set of 11 candidate 22q13.3 PMS modifier genes assembled from the existing studies based on pLI (>0.9), population impact factor (>0.5), and functional evidence for neurodevelopmental or motor phenotypes.^17,18,30,31^ Genomic coordinates for candidate genes were obtained from GENCODE v44. A deletion was considered to involve a candidate gene if it overlapped any portion of the gene body. Deletions were stratified into size tertiles (Small < 0.10 Mb, Medium 0.10–0.75 Mb, Large > 0.75 Mb) for visualization.

### Statistical Analysis of Phenotypic Differences

Phenotypic comparisons were conducted using non-parametric tests given the non-normal distributions of most phenotypic variables. All analyses were performed in R.

First, we compared all *SHANK3* variant carriers combined (PTV, deletion, missense, and duplication) against non-carrier ASD cases using Wilcoxon rank-sum tests for each phenotypic variable. Second, we performed separate comparisons for each variant type (PTV, deletion, missense, and duplication) against non-carrier ASD cases. Third, pairwise comparisons between all variant types were conducted using Wilcoxon rank-sum tests. Fourth, Kruskal-Wallis tests were used as omnibus tests to assess whether phenotypic measures differed across the four variant types. For all Wilcoxon rank-sum tests, the rank-biserial correlation (r) was calculated as a measure of effect size, where values closer to 1 or -1 indicate larger group differences. *P* values were corrected for multiple testing using the Benjamini-Hochberg (BH) method within each comparison group.

To focus on damaging variants, we performed a secondary analysis excluding duplication carriers, comparing the remaining *SHANK3* damaging variant carriers (PTV, deletion, and missense combined) against non-carrier ASD cases. Results from this analysis were visualized using a volcano plot displaying effect sizes (rank-biserial r) against BH-adjusted *P* values, and a heatmap summarizing signed effect sizes across variant types relative to non-carrier ASD cases.

### Phenotypic Threshold Sensitivity Analysis

To explore whether combinations of phenotypic measures could discriminate *SHANK3* damaging variant carriers (PTV, deletion, and pathogenic missense) from non-carrier ASD cases, we conducted a systematic two-measure threshold sensitivity analysis. We selected 16 phenotypic variables that showed significant associations with *SHANK3* damaging status, including cognitive ability, motor coordination (DCDQ total and subscale scores), repetitive behaviors (RBS-R total and subscale scores), and developmental milestones (ages at walking, self-feeding with a spoon, and combining phrases). For each variable, a set of candidate thresholds was defined based on clinically meaningful ranges, with the direction of impairment specified a priori (e.g., cognitive scores below threshold, milestone ages above threshold).

All possible pairwise combinations of the 16 measures were tested. For each pair, only individuals with non-missing data on both measures (complete cases) were included. At each combination of thresholds, sensitivity was calculated as the proportion of *SHANK3* damaging variant carriers meeting both criteria, and specificity as the proportion of non-carrier ASD cases not meeting both criteria. The Youden index (sensitivity + specificity - 1) was used to rank threshold combinations. Results were filtered by minimum sample size (n >= 10 *SHANK3* damaging variant carriers) and minimum specificity (> 0.80) to identify clinically viable operating points.

Based on the systematic sweep, cognitive ability (IQ < 70) and motor coordination (DCDQ total score < 35) were identified as a high-performing two-measure combination. To evaluate the discriminative performance of these measures individually and in combination, receiver operating characteristic (ROC) curves were generated using logistic regression. Three models were compared: IQ alone, DCDQ alone, and IQ combined with DCDQ, with the area under the curve (AUC) computed for each. Enrichment of the combined phenotypic profile among *SHANK3* damaging variant carriers relative to non-carrier ASD cases was assessed using Fisher’s exact test, and the odds ratio with 95% confidence interval was reported.

### Gene-level Enrichment and Pathway Analysis in *SHANK3*-phenotype Cases

To investigate whether ASD cases meeting the *SHANK3*-associated phenotypic profile (IQ < 70 and DCDQ total score < 35) harbor an excess of rare deleterious variants in specific genes, we performed a gene-level enrichment analysis comparing 1,421 *SHANK3*-phenotype cases against 7,871 non-phenotype ASD cases (those not meeting both criteria, with non-missing data on both measures). Rare deleterious variants included de novo PTVs, genome-wide rare PTVs, and rare deletions. Missense variants and duplications were excluded from this analysis.

For CNVs spanning multiple genes, each overlapping gene was considered individually. All variant sources were restricted to 255 ASD risk genes with false discovery rate < 0.1 from the Transmission and De novo Association (TADA) analysis in the Fu et al., 2022^16^.

For each gene, the number of unique individuals carrying at least one rare deleterious variant was counted in both the *SHANK3*-phenotype and non-phenotype groups. Enrichment was assessed using two-sided Fisher’s exact tests, and odds ratios with *P* values were computed for each gene. *P* values were corrected for multiple testing using the Benjamini-Hochberg method.

Genes nominally enriched in the *SHANK3*-phenotype group (*P* < 0.05, odds ratio > 1) were submitted to pathway enrichment analysis using EnrichR. Multiple pathway databases were queried, including KEGG 2021^28^, Reactome 2022^29^, Gene Ontology (Biological Process 2023)^27^, and SynGO 2024^26^. Results were ranked by adjusted *P* value, and pathways with Benjamini-Hochberg adjusted *P* < 0.05 were reported.

### Quality control of genome-wide association study data

Genotype data from the SPARK cohort were generated using two platforms across nine batches: batches 1–4 (v1: 27,120 samples and 634,851 variants; v2: 42,981 samples and 649,528 variants) were genotyped on the Illumina Global Screening Array (GSA), while batches 5–9 (71,267 samples and 1,414,695 variants) were genotyped using TWIST genotyping-by-sequencing (GxS). We also utilized the Mount Sinai Million (MSM) GxS data including 28,227 samples and 1,414,695 variants. Due to differences in variant coverage and genotyping technology, quality control (QC) was performed separately for each platform and performed again after merging all datasets using shared variants following established guidelines for GWAS in ancestrally diverse populations^46^.

Standard sample-level QC included removal of individuals with genotype missingness rate > 2%, excess autosomal heterozygosity (|F| > 0.2), and discordance between reported and genetically inferred sex. Variant-level QC removed single-nucleotide polymorphisms (SNPs) with missingness rate > 2%, minor allele frequency (MAF) < 1%, and significant deviation from Hardy-Weinberg equilibrium (HWE) test (*P* < 1 × 10^-5^).

To minimize confounding from population stratification, we adopted ancestry-stratified quality control approach. Principal component analysis (PCA) was performed by merging QC-passed samples with the 1000 Genomes Project Phase 3 reference panel^47^. Samples were assigned to five major ancestry groups based on their proximity to reference populations: non-Finnish European (NFE), Admixed American (AMR), African (AFR), East Asian (EAS), and South Asian (SAS) using a random forest classifier trained on 1000 Genomes Project reference populations. Ancestry-specific QC, including heterozygosity filtering and HWE testing, was subsequently performed within each ancestry group.

Haplotype phasing was performed using SHAPEIT v5.1.0^48^, incorporating pedigree information to improve phasing accuracy in family-based samples. Genotype imputation was conducted using IMPUTE5 v1.2.0 with the 1000 Genomes Project Phase 3 as the reference panel^49^. To ensure unbiased estimation of imputation quality in the presence of related individuals, INFO scores were calculated using founders only. Post-imputation QC retained variants with INFO score > 0.8 and MAF > 1%.

### Polygenic risk score calculation

To assess polygenic burden for psychiatric disorders, we calculated PRSs for ASD, ADHD, OCD, and schizophrenia using publicly available GWAS summary statistics ^50–53^. For PRS comparisons, ASD cases were divided into two groups based on the phenotypic criteria defined above: *SHANK3*-phenotype cases (IQ < 70 and DCDQ < 35) and non-*SHANK3*-phenotype cases (not meeting both criteria), regardless of *SHANK3* variant carrier status. The MSM samples without mental disorders served as the control group. To avoid population stratification bias, PRS analyses were restricted to individuals of non-Finnish European (NFE) ancestry, involving 753 *SHANK3*-phenotype ASD cases, 5,192 non-*SHANK3*-phenotype ASD cases, and 8,198 unaffected controls.

PRSs were computed using LDpred2 ^54^, a Bayesian method that accounts for linkage disequilibrium (LD) between variants and provides improved prediction accuracy compared to traditional clumping and thresholding approaches. LD reference panels were constructed from a random subset of unrelated NFE individuals in the SPARK cohort. For each trait, we applied the LDpred2-auto model, which automatically estimates the optimal SNP heritability and polygenicity parameters from the data without requiring hyperparameter tuning. Variants were filtered to HapMap3 SNPs to ensure robust LD estimation ^55^. PRSs were standardized to have mean zero and unit variance within the analytic sample.

## Acknowledgements

We are grateful to all of the families in SPARK, the SPARK clinical sites and SPARK staff.

We acknowledge the Mount Sinai Million Health Discoveries Program at the Charles Bronfman Institute for Personalized Medicine, Icahn School of Medicine at Mount Sinai, for providing access to data resources used in this study. The Mount Sinai Million Health Discoveries Program is an initiative integrating genomic, health, and research data from Mount Sinai patients to advance discovery across diverse populations.

This study was supported by a grant from the Beatrice and Samuel A. Seaver Foundation (BM) and the National Institute of Mental Health (NIMH; R01MH139952).

## Author contributions

Study concept and design: A.K., S.J., B.M.

Acquisition, analysis, or interpretation of data: All authors

Drafting of the manuscript: A.U, S.S., M.C., A.R., S.J, B.M

Critical revision of the manuscript for important intellectual content: All authors

Statistical analysis: A.U, S.J, B.M

Obtained funding: B.M

Study supervision: S.J, B.M

## Competing interests

The authors declare no competing interests.

## Data availability

The data that support the findings of this study are available from SPARK and the Mount Sinai Million Health Discoveries Program, but restrictions apply to the availability of these data, which were used under license for the current study and are therefore not publicly available. Data are available from the corresponding author upon reasonable request and with permission of the respective data providers.

## Code availability

Code and resources used in this study is available on GitHub at (https://github.com/MahjaniLab/shank3_rare). For any further inquiries or requests for code not available in the repository, please contact the corresponding author.

## Notes

### Competing Interest Statement

The authors have declared no competing interest.

